# Molecular surveillance of multidrug-resistant tuberculosis at the dawn of the genomic era, Argentina, 2013–2022

**DOI:** 10.64898/2026.01.27.26344616

**Authors:** Federico Lorenzo, Roxana Paul, Johana Monteserin, Néstor Masciotra, Eduardo Mazzeo, Ingrid Wainmayer, Laura Pérez-Lago, Mario Matteo, Ana Gamberale, Hugo Fernández, Lucía Elvira Irazu, Marcelo Adrián Rodríguez, Domingo Palmero, Darío García de Viedma, Norberto Simboli, Beatriz López, Noemí Kaoru Yokobori

## Abstract

We genotyped 1189 multidrug-resistant *Mycobacterium tuberculosis* isolates identified during 2013–2022 in Argentina, through a mixed strategy using PCR-based methods and whole-genome sequencing. Epidemiological, geographic distribution and microbiological data were integrated. Most cases belonged to a cluster and orphan cases were associated with history of poor adherence to treatment and foreign-born patients. The Euro-American lineage4 was predominant. The most important clusters, M, Ra, Rb and Callao2 strains, comprised 45.9% of the newly diagnosed cases. The historically most important M strain started to decline. The Rb strain was found to be geographically widespread and was strongly associated with transgender women. The Callao2 strain, probably imported from Peru, expanded significantly in the latest years. A preliminary genomic analysis identified several possible local transmission chains. The relatively low proportion of preXDR/XDR cases is encouraging but deserves close monitoring. Collectively, our results constitute important groundwork for the full transition to WGS-based surveillance.

**One-sentence summary:** We evaluated the genotypes associated with multidrug-resistant tuberculosis in Argentina, 2013–2022.

## Introduction

Multidrug resistance (MDR) is a major challenge faced by global tuberculosis control. Due to its costly treatment, low success rate and higher mortality, the epidemiological surveillance of MDR-tuberculosis cases is crucial. Resistance can emerge from the selection of mutant clones under intermittent treatment or can be due to the transmission of prevalent MDR clones. Each situation requires specific interventions, such as closer treatment monitoring and thorough contact tracing.

Argentina has a moderate tuberculosis burden, but an alarming rise has been observed in the recent years from 21.3/100,000 population in 2013 to 35.4/100,000 in 2024 (1,2). The proportion of MDR cases remained relatively stable over the years, representing 1 – 2% of all the tuberculosis cases (3). In Argentina, MDR-tuberculosis emerged in the 1990’s in consonance with the HIV/AIDS pandemic, driven by epidemic clones related to discrete social and geographic niches (4–7). Systematic surveillance started in 2003 (6) and followed the evolution of molecular genotyping methods (8). Previous reports showed that clustering rate was high (75%) and the most important clones, the M, Ra and Rb strains, belonged to the Euro-American lineage, were autochthonous and no prior evidence showed a local expansion of imported strains (6). These strains emerged as hospital outbreaks in the cities of Buenos Aires (M strain)(7) and Rosario (Ra and Rb strains)(4) and spread to the community (9).

To evaluate the recent epidemiological landscape of MDR tuberculosis in Argentina, herein, we analyzed the MDR-*Mycobacterium tuberculosis* genotypes identified during the 2013 – 2022 period through a mixed strategy. We contrasted the cases due to clustered vs. orphan genotypes, the latest trends among the most important clones, and resistance to second-line drugs.

## Methods

### Isolate selection and genotyping

The Mycobacteria Service is the National Reference Laboratory for the diagnosis of tuberculosis and systematically surveys MDR-tuberculosis cases since 2003. Figure 1 shows the inclusion/exclusion criteria and study design. In the 2013–2022 period, 1189 MDR *M. tuberculosis* isolates were received, representing MDR cases diagnosed nationwide (Appendix 1). Among these, 1169 isolates were genotyped using three genotyping methods depending on their availability across the study period; a tailored multiplex PCR named TRAP for the detection of the M, Ra and Rb strains (2015–2020)(9,10), 15 loci mycobacterial interspersed repetitive-unit-variable-number tandem-repeat (MIRU-VNTR; 2013–2020)(11) or whole genome sequencing (WGS; 2021 onwards) were applied (Figure 1). Some older isolates had been sequenced for prior research projects (9,10,12). We performed validation assays in each methodological transition, and a consolidated classification criterion was applied. A single nucleotide polymorphism (SNP) distance of ≤20 among monophyletic isolates was considered for WGS-based genotyping for a better concordance with MIRU-VNTR (Appendix 1, Appendix 2). MIRU-VNTR types were analyzed in the *MIRU-VNTRplus* platform (13) and *Run TB-lineage* was used to infer their lineage (14). The selected isolates corresponded to 955 patients, of which 910 were diagnosed with MDR-tuberculosis during the study period. The latter group was considered for demographic and clustering analysis. Some patients had more than one isolate sent to our laboratory (Figure 1).

**Figure 1.**
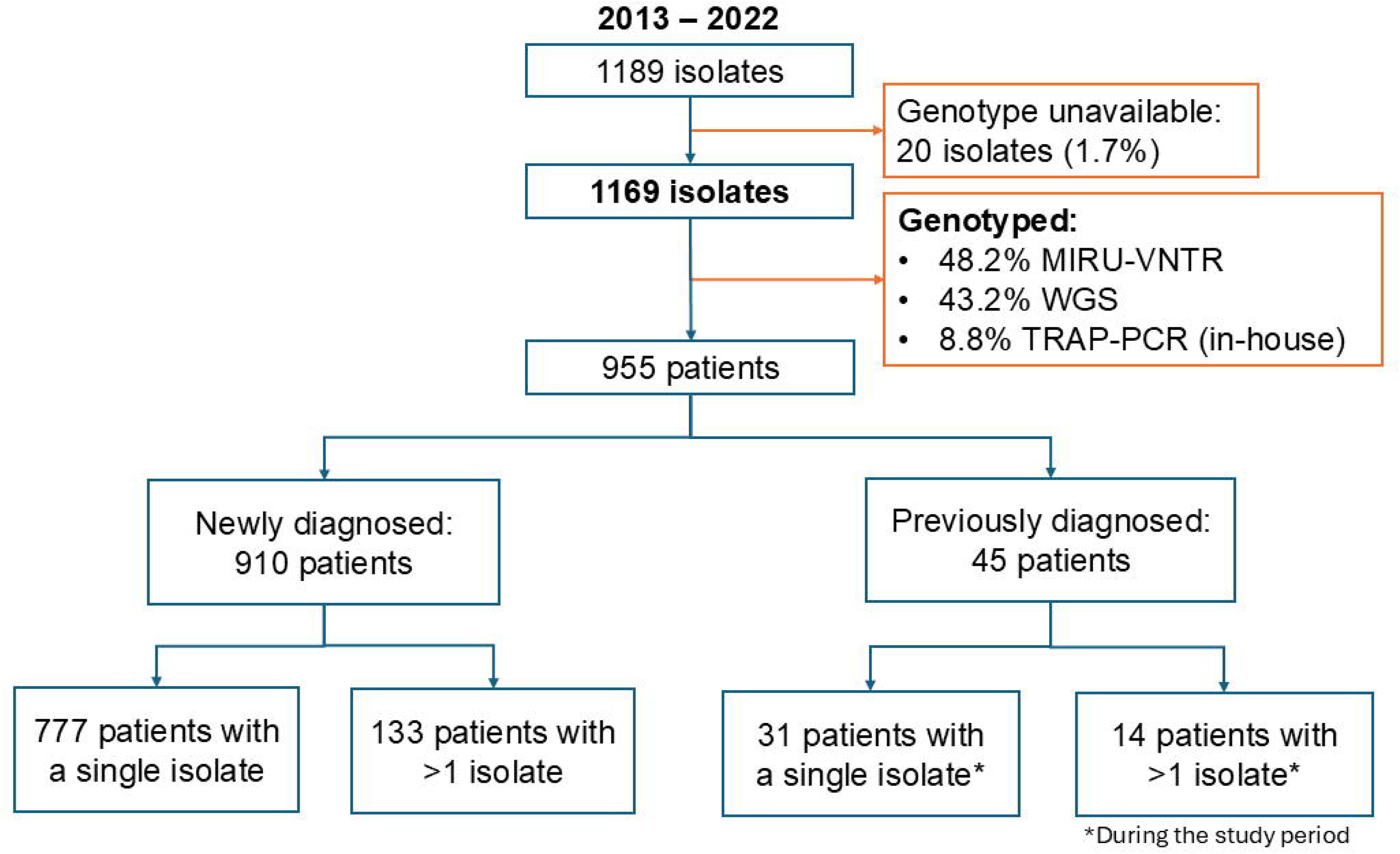
Inclusion and exclusion criteria of MDR *M. tuberculosis* isolates that were sent to the *Servicio de Micobacterias*, the National Reference Laboratory of Argentina, in the 2013-2022 period. Isolates that could not be genotyped or had mixed clones were excluded. The remaining isolates were genotyped using a combined strategy (Appendix 1). An in-house test named MAS-PCR, which detects the most important resistance conferring mutations for rifampicin and isoniazid, was applied to all the isolates upon reception. Genotyping results were integrated with microbiological and epidemiological data as detailed in the Materials and Methods section and Appendix 1. Patients were classified according to their date of MDR-tuberculosis diagnosis and the number of isolates sent to our laboratory during the study period. MIRU-VNTR: mycobacterial interspersed repetitive-unit-variable-number tandem-repeat; WGS: whole genome-sequencing; TRAP-PCR: in-house PCR for the detection of M, Ra and Rb strains.

For WGS analysis, DNA was recovered from Löwenstein-Jensen slants using the cetyltrimethylammonium bromide method (15). Genomic libraries were prepared and indexed using the *Preparation* and the *Barcoding* kits (Illumina), following the instructions of the manufacturer, and sequenced in the Illumina MiSeq or Novaseq platform at the UOCNGyB, ANLIS, with paired-end mode. Quality was evaluated using *FastQC* v0.12.1. The average sequencing depth was 80x. Reads were aligned to the H37Rv genome using *BWA* 0.7.17 (16). Allelic variants were called with *GATK* 4.4.0.0 (17) and were annotated with *snippy* (18). Due to uncertainty in their alignments, variants in PE/PPE genes, repetitive regions and mobile elements were excluded (19,20). Maximum likelihood phylogenetic trees were constructed with *RAxML* (21) and visualized with *Microreact* (22). Spoligotypes and lineages were retrieved in silico using *TB-profiler* (23). Raw sequences were deposited in the NCBI (Appendix 3).

### Phenotypic and molecular drug-susceptibility testing

Routinely, *M. tuberculosis* complex isolates suspected of drug-resistance were evaluated with an in-house PCR-based test named MAS-PCR (24), which targets the wild type *rpo*B (codons 452, 450, 445 and 435), *fab*G1-*inh*A (−15 position) and *kat*G (codon 315) genes to infer the resistance to rifampicin (RIF) and isoniazid (INH), and were tested with phenotypic methods at the time of reception to resolve discordant results. Across the study period, the critical concentrations for the different drugs were adapted to the international guidelines. As a reference, in the latest years, the following critical concentrations were used for MGIT960-based testing: RIF (0.5 mg/L), INH (0.1 mg/L), ethambutol (EMB; 5 mg/L), ethionamide (ETH; 5 mg/L), kanamycin (2.5 mg/L), capreomycin (2.5 mg/L), amikacin (AMK; 1 mg/L), levofloxacin (1 mg/L), clofazimine (1 mg/L), linezolid (LZD; 1 mg/L), moxifloxacin (MFX) at low (0.25 mg/L) and high concentrations (1 mg/L) and bedaquiline (BDQ; 1 mg/L). Resistance to EMB (2 mg/L), ETH (40 mg/L) and capreomycin (40 mg/L) were also assessed by the proportion method in Löwenstein-Jensen medium. Resistance conferring mutations in the available genomes were detected using *TB-profiler* v6.2 (23) and were contrasted with the WHO catalogue of mutations v2 (25). PreXDR cases were defined as MDR isolates with resistance to at least a fluoroquinolone (FLQ) and XDR as cases with additional resistance to a group A drug (BDQ or LNZ) (26). Initial and final drug-resistance status were registered.

### Data processing and statistical analysis

The Committee in Research Ethics of the INE “Dr. Jara”, ANLIS, Argentina approved the project and waived the informed consent requirement (YOKOBORI05/2022). Epidemiological, microbiological and genotyping information of each isolate was stored in a tailored database. Data were analyzed and visualized using the *tidyverse* (v2.0.0) and *base* packages in R (v4.5.1). Geographic distribution was processed and visualized using *sf* and *rnaturalearth* packages. Univariate statistical analyses of continuous variables were performed with Kruskal-Wallis test followed by Wilcoxon rank-sum test. Chi-squared test or Fisher’s exact test were used for categorical variables. Multinomial logistic regression with backward stepwise selection of variables was applied (Appendix 4), based on Akaike Information Criteria. For corrected analysis of time series, the Cochran-Armitage method was used. Statistical significance was considered for a p-value <0.05 for all the tests, with multiple logistic regressions additionally requiring 95% confidence intervals (CI) that excluded 1 for the identification of independent associations.

## Results

### General characteristics and geographic distribution of the cases

We evaluated the genotypes of 1169 MDR *M. tuberculosis* isolates sent to the National Reference Laboratory during 2013–2022 (mean ± standard error of the mean: 117 ± 4.5 isolates/year). A total of 910 patients were newly diagnosed (Figure 1), which represents 80% of the notified cases for the same period (Appendix 1). The most frequent resistance mutations were in the positions *rpo*B450 (71.9%), *rpo*B435 (15.7%) and *rpo*B445 (4.5%) for RIF, and *katG*315 (66.9%) and -15_*fab*G_*inh*A (20.5%) for INH.

Among newly diagnosed MDR-tuberculosis cases, working-age males were the most represented (Table 1). Foreign-born patients constituted 22.3%, among which patients born in Peru, the South American country with the highest DR rates (3), were the most represented. At least one risk factor for tuberculosis was registered for most cases (70%; Table 1). Transgender women were overrepresented (4.3%) compared to the general population of Argentina (0.13%)(27) and 74.4% of them were living with HIV.

**Table 1.**
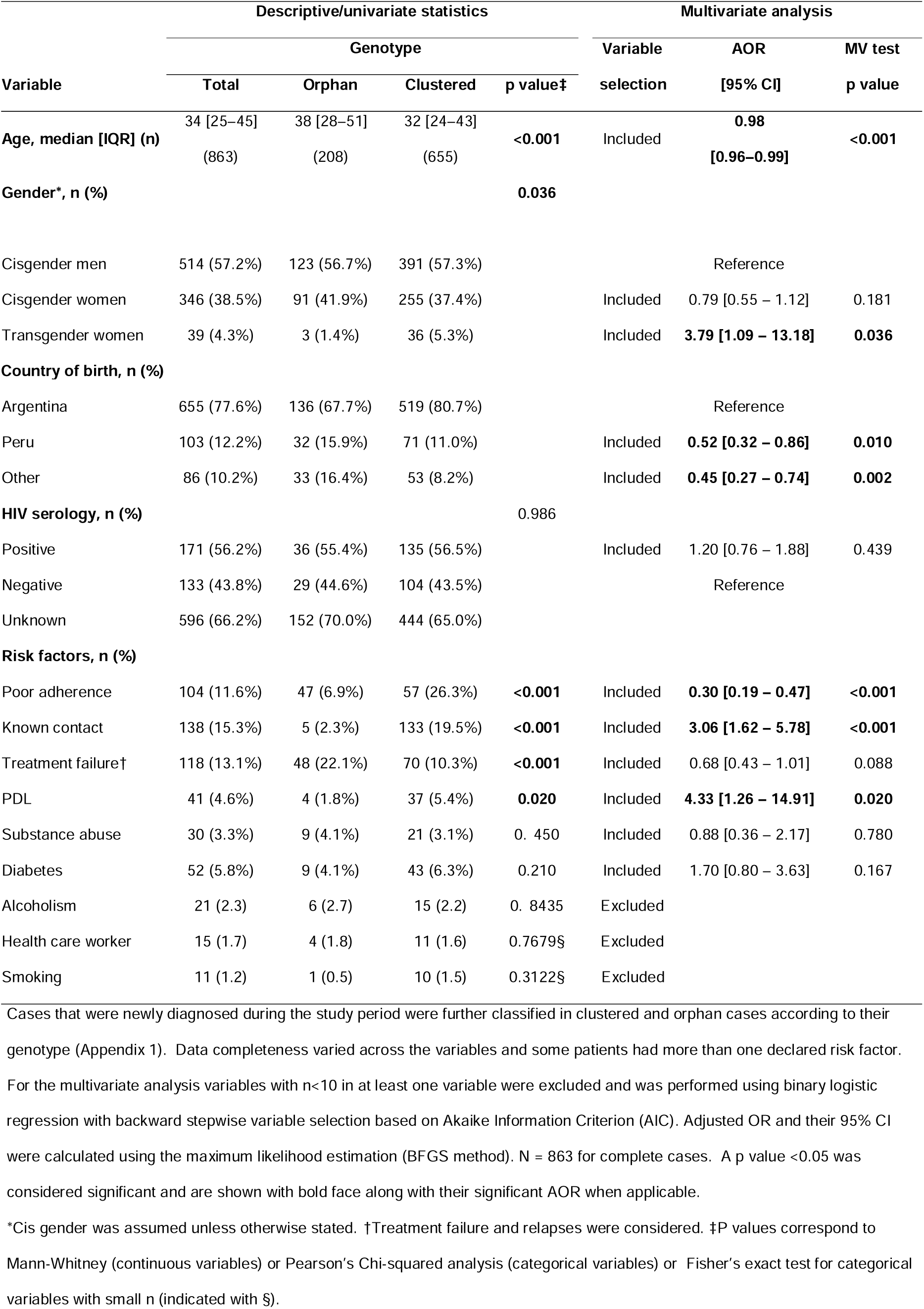
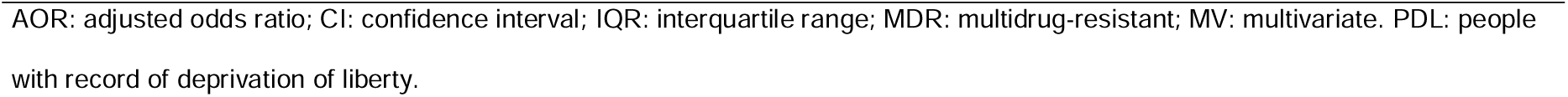
Descriptive statistics and multivariate analysis of the variables modeled in newly diagnosed patients affected by MDR *Mycobacterium tuberculosis* according to their clustering status, Argentina, 2013-2022.

| Variable | Descriptive/univariate statistics |  |  |  | Multivariate analysis |  |  |
| --- | --- | --- | --- | --- | --- | --- | --- |
|  | Genotype |  |  |  | Variable selection | AOR<br>[95% CI] | MV test<br>p value |
|  | Total | Orphan | Clustered | p value‡ |  |  |  |
| **Age, median [IQR] (n)** | 34 [25–45]<br>(863) | 38 [28–51]<br>(208) | 32 [24–43]<br>(655) | **<0.001** | Included | **0.98**<br>**[0.96–0.99]** | **<0.001** |
| **Gender\*, n (%)** |  |  |  | **0.036** |  |  |  |
| Cisgender men | 514 (57.2%) | 123 (56.7%) | 391 (57.3%) |  |  | Reference |  |
| Cisgender women | 346 (38.5%) | 91 (41.9%) | 255 (37.4%) |  | Included | 0.79 [0.55 – 1.12] | 0.181 |
| Transgender women | 39 (4.3%) | 3 (1.4%) | 36 (5.3%) |  | Included | **3.79 [1.09 – 13.18]** | **0.036** |
| **Country of birth, n (%)** |  |  |  |  |  |  |  |
| Argentina | 655 (77.6%) | 136 (67.7%) | 519 (80.7%) |  |  | Reference |  |
| Peru | 103 (12.2%) | 32 (15.9%) | 71 (11.0%) |  | Included | **0.52 [0.32 – 0.86]** | **0.010** |
| Other | 86 (10.2%) | 33 (16.4%) | 53 (8.2%) |  | Included | **0.45 [0.27 – 0.74]** | **0.002** |
| **HIV serology, n (%)** |  |  |  | 0.986 |  |  |  |
| Positive | 171 (56.2%) | 36 (55.4%) | 135 (56.5%) |  | Included | 1.20 [0.76 – 1.88] | 0.439 |
| Negative | 133 (43.8%) | 29 (44.6%) | 104 (43.5%) |  |  | Reference |  |
| Unknown | 596 (66.2%) | 152 (70.0%) | 444 (65.0%) |  |  |  |  |
| **Risk factors, n (%)** |  |  |  |  |  |  |  |
| Poor adherence | 104 (11.6%) | 47 (6.9%) | 57 (26.3%) | **<0.001** | Included | **0.30 [0.19 – 0.47]** | **<0.001** |
| Known contact | 138 (15.3%) | 5 (2.3%) | 133 (19.5%) | **<0.001** | Included | **3.06 [1.62 – 5.78]** | **<0.001** |
| Treatment failure† | 118 (13.1%) | 48 (22.1%) | 70 (10.3%) | **<0.001** | Included | 0.68 [0.43 – 1.01] | 0.088 |
| PDL | 41 (4.6%) | 4 (1.8%) | 37 (5.4%) | **0.020** | Included | **4.33 [1.26 – 14.91]** | **0.020** |
| Substance abuse | 30 (3.3%) | 9 (4.1%) | 21 (3.1%) | 0.450 | Included | 0.88 [0.36 – 2.17] | 0.780 |
| Diabetes | 52 (5.8%) | 9 (4.1%) | 43 (6.3%) | 0.210 | Included | 1.70 [0.80 – 3.63] | 0.167 |
| Alcoholism | 21 (2.3) | 6 (2.7) | 15 (2.2) | 0.8435 | Excluded |  |  |
| Health care worker | 15 (1.7) | 4 (1.8) | 11 (1.6) | 0.7679§ | Excluded |  |  |
| Smoking | 11 (1.2) | 1 (0.5) | 10 (1.5) | 0.3122§ | Excluded |  |  |

Cases that were newly diagnosed during the study period were further classified in clustered and orphan cases according to their genotype (Appendix 1). Data completeness varied across the variables and some patients had more than one declared risk factor.
For the multivariate analysis variables with n<10 in at least one variable were excluded and was performed using binary logistic regression with backward stepwise variable selection based on Akaike Information Criterion (AIC). Adjusted OR and their 95% CI were calculated using the maximum likelihood estimation (BFGS method). N = 863 for complete cases. A p value <0.05 was considered significant and are shown with bold face along with their significant AOR when applicable.
\*Cis gender was assumed unless otherwise stated. †Treatment failure and relapses were considered. ‡P values correspond to Mann-Whitney (continuous variables) or Pearson's Chi-squared analysis (categorical variables) or Fisher's exact test for categorical variables with small n (indicated with §).
AOR: adjusted odds ratio; CI: confidence interval; IQR: interquartile range; MDR: multidrug-resistant; MV: multivariate. PDL: people with record of deprivation of liberty.

A higher incidence of MDR-tuberculosis cases was observed in Buenos Aires province and Buenos Aires Autonomous City, followed by Santa Fe province (Table 2), especially in their densely populated urban areas (data not shown).

**Table 2.** Geographic distribution of newly diagnosed MDR-tuberculosis cases, Argentina, 2013-2022.

| Region; n = 839 | Subtotal (%) | Genotype |  |
| --- | --- | --- | --- |
|  |  | Clustered | Orphan |
| Buenos Aires province | 368 (43.9) | 251 (69.3) | 117 (31.8)* |
| Buenos Aires autonomous city | 187 (22.3) | 146 (78.1) | 41 (21.9) |
| Santa Fe province | 127 (15.1) | 122 (96.1)* | 5 (3.9) |
| Northwestern provinces | 78 (9.3) | 64 (82.0) | 14 (18.0) |
| Other provinces | 79 (9.4) | 50 (63.3) | 29 (36.7) |

\*Standardized residual value that were higher than the critical value. MDR: multidrug resistance.
Cases were grouped considering their declared place of residence. The northwestern provinces of Tucumán, Salta and Jujuy were grouped. Thirty-one cases corresponding to persons deprived of liberty were excluded because their primary place of residence was not declared. Clustered vs. orphan cases were analyzed using a Chi-squared test; $p < 0.001$ . Standardized residuals analysis was used as post-hoc test, considering a critical value of 2.81 using Bonferroni correction.

### Clusters and orphan genotypes

Of the newly diagnosed cases, 75.9% belonged to a cluster. Cases due to orphan genotypes were associated with higher age, foreign born patients and history of poor adherence to treatment, while clustered cases were associated with transgender women, patients with a known contact and persons deprived of liberty (PDL; Table 1). Clustering rates were significantly higher in Santa Fe province, mainly in Rosario city (92.9% of the cases diagnosed in Santa Fe). Conversely, Buenos Aires province had more orphan cases than expected (Table 2).

The Euro-American lineage was virtually predominant, in line with other reports from the region (28,29). East Asian isolates were rare and were associated with migrants from regions with high prevalence (Appendix 5) including Peru (30,31). The most frequently found WGS-based lineages were 4.3.3, 4.3.2 and 4.1.2.1 (T1, LAM3 and, LAM9 spoligotypes). Orphan isolates showed greater clade diversity (Appendix 5).

A total of 48 clusters were identified. The median number of isolates from newly diagnosed cases per cluster was three (interquartile range [IQR]: 2–8). Five clusters belonged to the top decile according to their size, with 70 or more isolates each (Appendix 6). Among them, the O genotype included at least three divergent genotypes that could not be fully classified with the current approach (Appendix 1 and 2). Hereafter, we will refer to the four remaining largest clusters as the major clusters of the period.

### Major clusters and the Callao2 outbreak

The newly diagnosed cases due to the four most important MDR clusters, the Rb, Ra, M and Callao2 strains, collectively represented 45.7% of all the cases, and were constitutively resistant to three or more drugs (Table 3).

**Table 3.** Characteristics of the major MDR *Mycobacterium tuberculosis* clusters circulating in Argentina, 2013-2022.

| Variable | Strain |  |  |  |
| --- | --- | --- | --- | --- |
|  | Callao2 | Rb | Ra | M |
| Genotype |  |  |  |  |
| Spoligotype (SIT) | LAM6 (1355) | T1 (159) | LAM3 (33) | H2 (2) |
| WGS lineage | 4.3.3 | 4.3.3 | 4.3.2 | 4.1.2.1.1 |
| DR profile* | INH, RIF, PZA,<br>EMB | INH, RIF, ETH | INH, RIF, PZA | INH, RIF, PZA,<br>EMB, STR,<br>AMK |
| Resistance conferring mutations* | *katG*_S315T,<br>*rpoB*_D435V,<br>*pncA*_Q10R,<br>*embB*_Y319S | *fabG1*_c-15t,<br>*rpoB*_S450L | *katG*_S315T,<br>*rpoB*_S450L,<br>*pncA*_S104R | *katG*_S315T,<br>*rpoB*_S450L,<br>*pncA*_Q10P,<br>*embB*_G406A,<br>*gidB*_V110fs,<br>*rrs*_a1401g |

\*The most frequently found drug-resistance profiles and mutations are indicated.
AMK: amikacin; DR: drug-resistance; EMB: ethambutol; ETH: ethionamide; INH: isoniazid;
PZA: pyrazinamide; RIF: rifampicin; SIT: spoligotype international type; STR: streptomycin.

Multinomial regression analysis considering Ra strain as the reference (Appendix 4) showed that the M strain was associated with history of treatment failure or relapse. Patients infected with the Rb strain were 14 times more likely to be transgender woman than those infected with the Ra strain. The Rb strain was also strongly associated with persons living with HIV or born in foreign countries including Peru. Patients infected with the Callao2 strain were younger and more likely to be born in Peru (Table 4). Some noticeable changes in their proportions were observed over the years. The Callao2 strain markedly expanded after 2018 (Figure 2; Cochran-Armitage z score: +5.316; p < 0.05), and newly diagnosed cases due to the M strain, which used to represent 29% of MDR cases in Argentina (6), declined (6.1 % in the 2021-2022 biennia, p<0.001; Figure 2). M and Ra strains were mainly found in their primary niches, Buenos Aires and Rosario city, Santa Fe province respectively, and sporadically in other provinces (Figure 3). Most cases due to the Rb strain were found in the metropolitan area of Buenos Aires, but it was also reported in 10 other provinces (Figure 3). Cases from the northwestern Tucumán, Salta and Jujuy provinces represented 9.6 % of the newly diagnosed cases due to the Rb strain, among whom 45.5 % were transgender women (data not shown).

**Figure 2.**
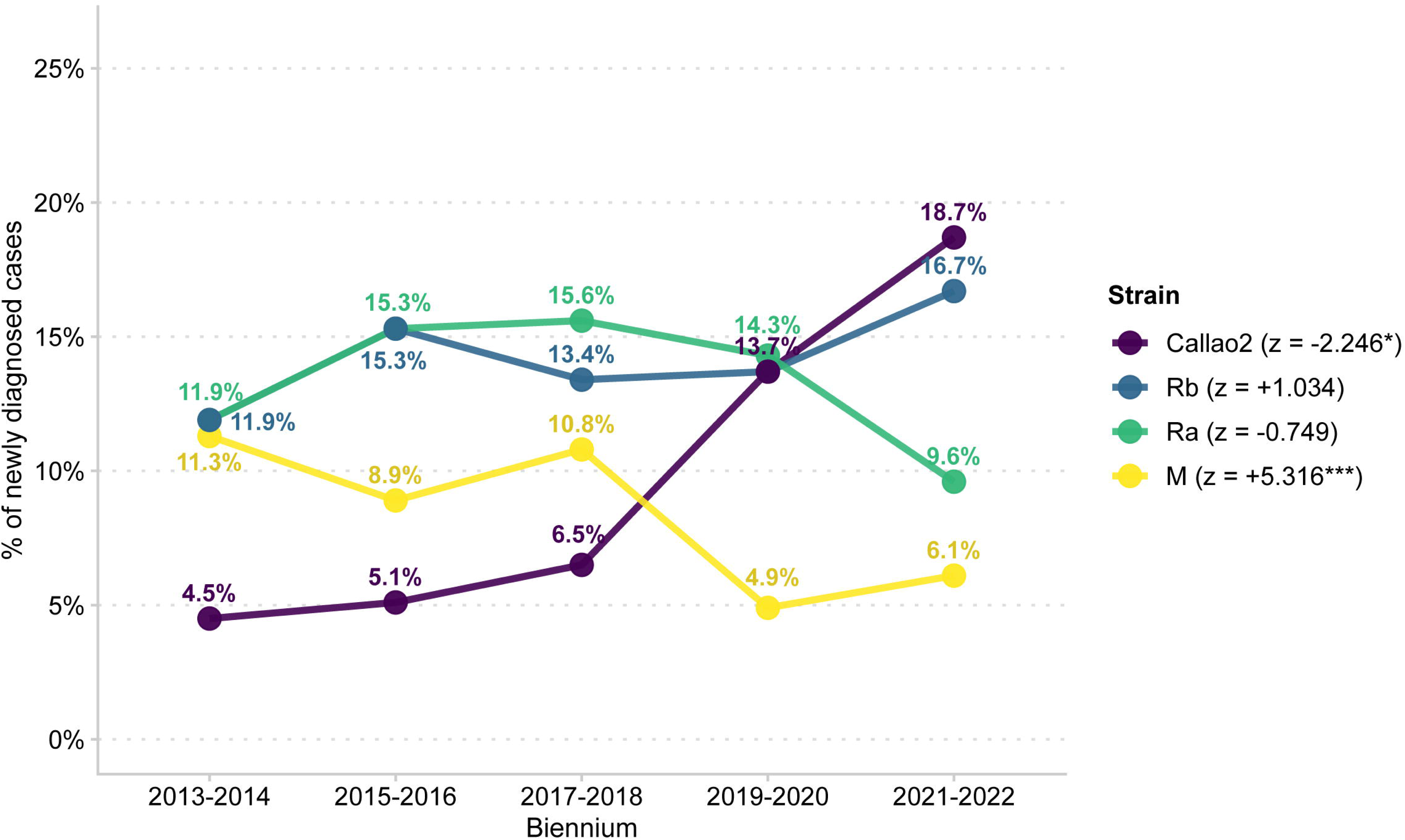
The percentage of cases due to the four major MDR-*M. tuberculosis* strains, Argentina, 2013-2022. The differences in percent points (relative variation) comparing the 2013 – 2014 vs. 2021 – 2022 biennia were of −5.2 (×0.54), −2.3 (×0.81), 4.8 (×1.4) and 14.2 (×4.16) for the M, Ra, Rb and Callao2 strains respectively. The bilateral Cochran-Armitage test with ordinal scores (0 – 4) was used to evaluate the lineal trend across the five biennia. The z-values are shown along with the legends. Global Chi-squared (four strains 5 biennia) = 37.730, degrees of freedom = 12, p < 0.001. p values < 0.05 were considered significant. *p < 0.05; ***p<0.001.

**Figure 3.**
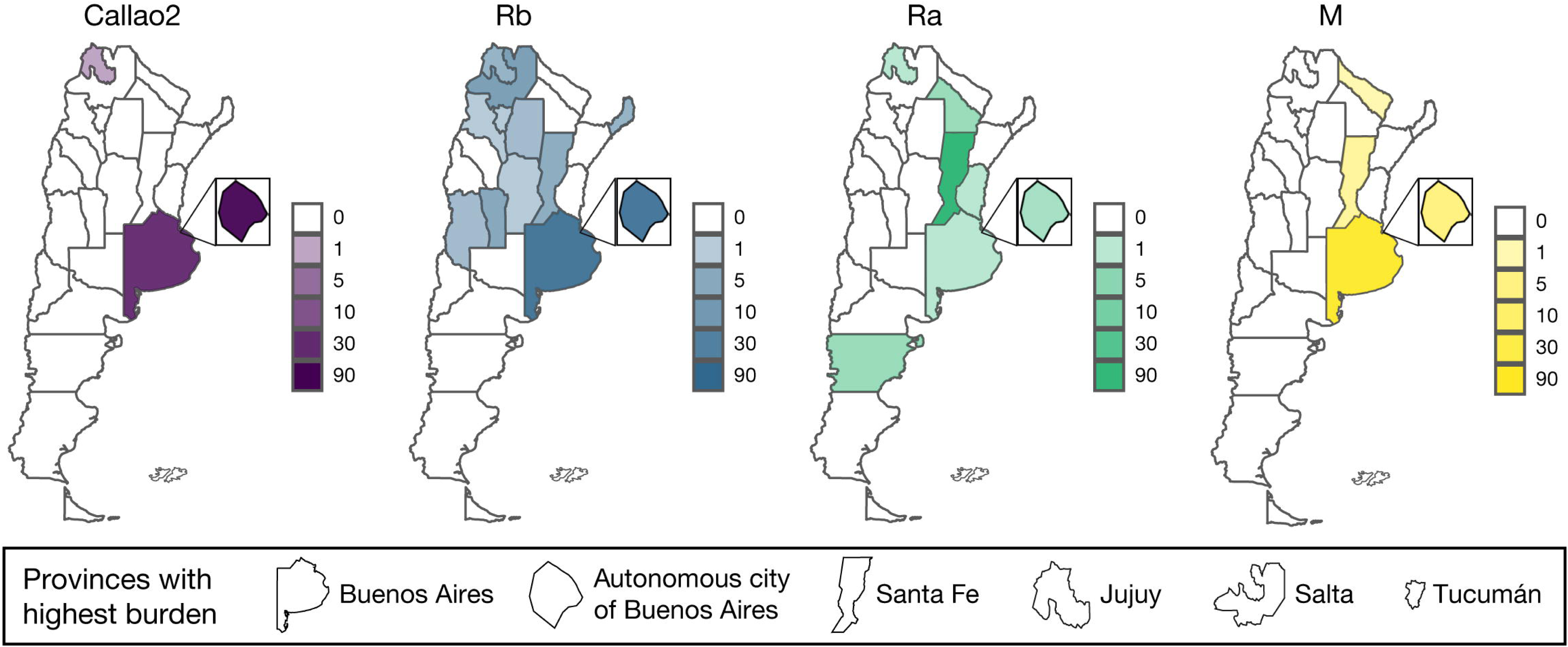
Provinces of Argentina showing the geographic distribution of newly diagnosed cases due to the Callao2, Rb, Ra and M strains. Darker colors indicate higher number of cases. Inset figure: amplification of the Autonomous City of Buenos Aires. The provinces with the highest burdens are indicated at the bottom of the figure.

**Table 4.**
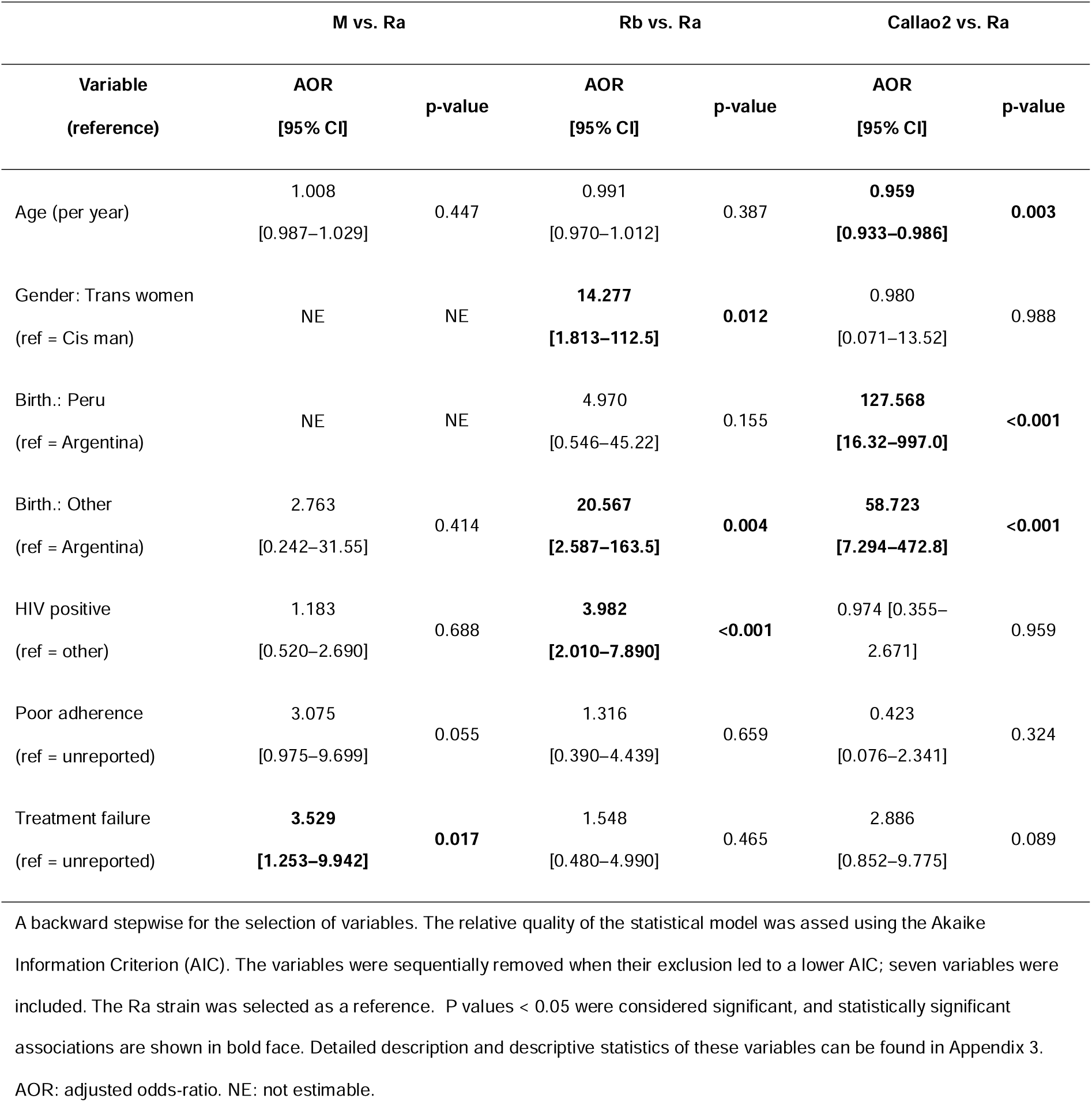
Multinomial logistic regression for the epidemiological variables and their associations with the four most important strains of the period.

|  | M vs. Ra |  | Rb vs. Ra |  | Callao2 vs. Ra |  |
| --- | --- | --- | --- | --- | --- | --- |
| Variable<br>(reference) | AOR<br>[95% CI] | p-value | AOR<br>[95% CI] | p-value | AOR<br>[95% CI] | p-value |
| Age (per year) | 1.008<br>[0.987–1.029] | 0.447 | 0.991<br>[0.970–1.012] | 0.387 | **0.959**<br>**[0.933–0.986]** | **0.003** |
| Gender: Trans women<br>(ref = Cis man) | NE | NE | **14.277**<br>**[1.813–112.5]** | **0.012** | 0.980<br>[0.071–13.52] | 0.988 |
| Birth.: Peru<br>(ref = Argentina) | NE | NE | 4.970<br>[0.546–45.22] | 0.155 | **127.568**<br>**[16.32–997.0]** | **<0.001** |
| Birth.: Other<br>(ref = Argentina) | 2.763<br>[0.242–31.55] | 0.414 | **20.567**<br>**[2.587–163.5]** | **0.004** | **58.723**<br>**[7.294–472.8]** | **<0.001** |
| HIV positive<br>(ref = other) | 1.183<br>[0.520–2.690] | 0.688 | **3.982**<br>**[2.010–7.890]** | **<0.001** | 0.974 [0.355–<br>2.671] | 0.959 |
| Poor adherence<br>(ref = unreported) | 3.075<br>[0.975–9.699] | 0.055 | 1.316<br>[0.390–4.439] | 0.659 | 0.423<br>[0.076–2.341] | 0.324 |
| Treatment failure<br>(ref = unreported) | **3.529**<br>**[1.253–9.942]** | **0.017** | 1.548<br>[0.480–4.990] | 0.465 | 2.886<br>[0.852–9.775] | 0.089 |

A backward stepwise for the selection of variables. The relative quality of the statistical model was assed using the Akaike Information Criterion (AIC). The variables were sequentially removed when their exclusion led to a lower AIC; seven variables were included. The Ra strain was selected as a reference. P values < 0.05 were considered significant, and statistically significant associations are shown in bold face. Detailed description and descriptive statistics of these variables can be found in Appendix 3. AOR: adjusted odds-ratio. NE: not estimable.

The steep growth of the Callao2 strain (Figure 2) suggested that a local outbreak took place in the metropolitan area of Buenos Aires. A phylogenetic analysis of the 61 available genomes of the Callao2 strain (68% of all the newly diagnosed cases) showed that this clade had a monophyletic origin (median of pairwise SNP distance [min–max]: 14 [0–33]) with three main branches (Figure 4). Recent transmission was suspected in at least five groups of patients considering a threshold of 5 SNPs (Appendix 7). The cluster A had the *rpo*B_S450L mutation and involved six patients born in Peru living in the metropolitan area of Buenos Aires (2 [0–4]; Figure 4). Among the isolates harboring the *rpo*B_D435V mutation, four subclusters suggested recent transmission. A cluster of six persons who cohabited with or were persons born in Peru (Figure 4, cluster B; 2 [0–7]), involved three preXDR cases sharing the *gyr*A_A90V substitution. A case of school transmission occurred in 2018 (Figure 4, cluster C). Patients in cluster D were mostly members of a native-born family living in a small informal settlement in Buenos Aires city (Figure 4; 1 [0–3]). The largest cluster included 37 patients diagnosed between 2019 and 2022 (Figure 4, cluster E; 1 [0–4]; 22 isolates with 0 SNPs of distance), who were born in Argentina (41.7%), Peru (30.6%) or Bolivia (25%). Patients were mainly residents of Buenos Aires city (69.4%), particularly of the adjacent neighborhoods of Flores and Floresta (15 patients), and informal settlements (10 patients). Two patients declared to work in sewing workshops. Most of these cases were diagnosed in 2021 (20 cases). A single patient was resident of Jujuy province who lived in Buenos Aires during the social isolation against the COVID-19 pandemic, declared in March 2020. Only 7 of these patients declared a direct epidemiological link to the outbreak.

**Figure 4.**
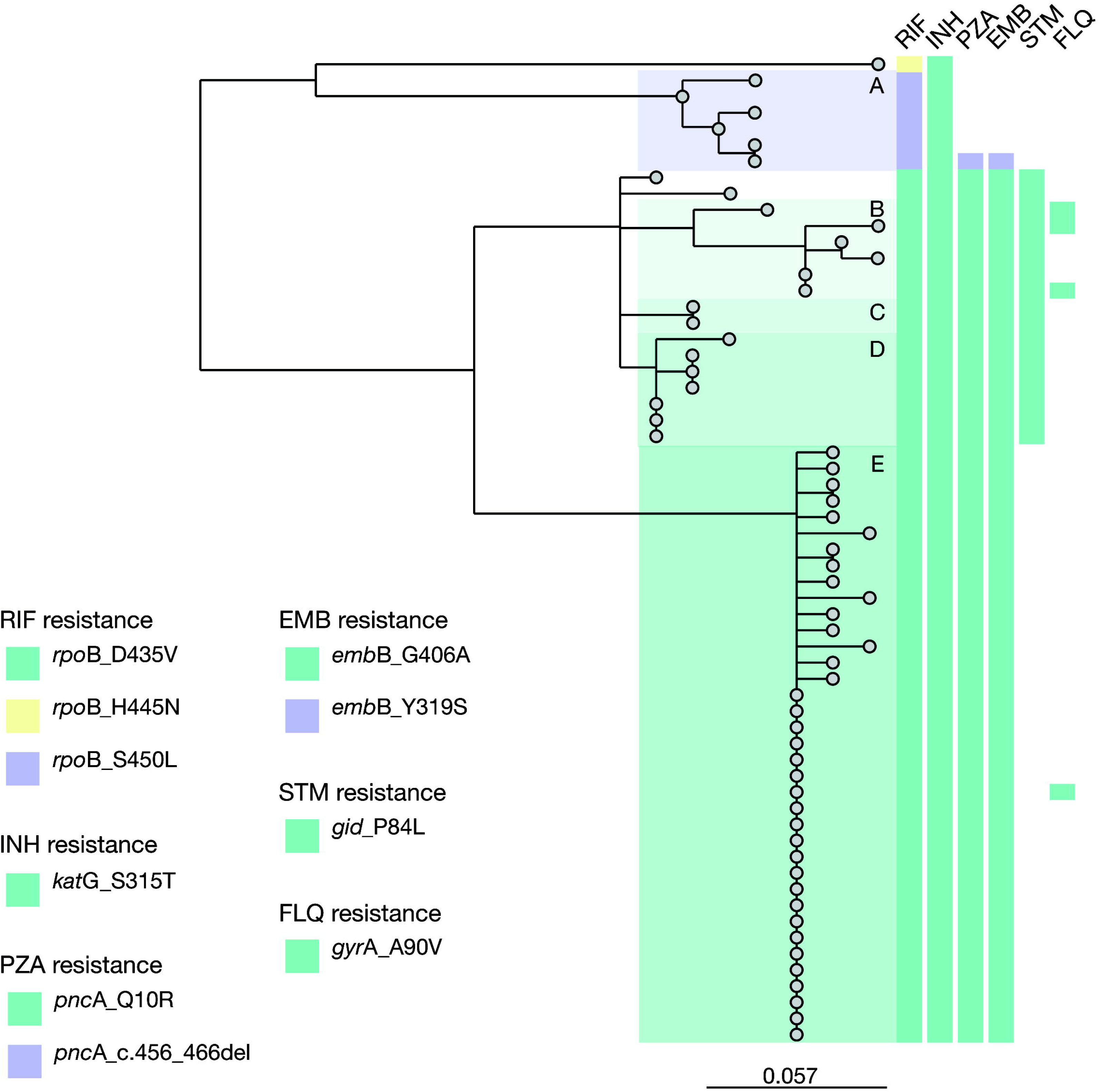
A maximum-likelihood phylogenetic tree of the 61 available genomes of Callao2 isolates, is shown. Blocks indicate the resistance-conferring mutations detected in each isolate, which are detailed in the legends. Isolates from newly diagnosed cases were included, except for one isolate from a patient who had been diagnosed 10 months earlier. The letters inside the shaded areas correspond to probable local transmission chains. **A.** Cluster sharing *rpo*B_S450L mutation, 2013 - 2022. **B.** Cluster including 3 preXDR cases, 2018 - 2022. **C.** School transmission, 2018. **D.** Intrafamilial transmission, 2013 - 2022. **E.** Largest cluster, 2019-2022. The scale bar indicates the number of substitutions per variable site. RIF: rifampicin; INH: isoniazid; PZA: pyrazinamide; EMB: ethambutol; STM: streptomycin; FLQ: fluoroquinolone.

### Minor clusters

The remaining 43 clusters were classified as minor clusters, of which 37 were identified in Buenos Aires area. Compared to the major clusters, patients in minor clusters were more frequently associated with a declared contact to an MDR-tuberculosis patient, mainly family members, and were less associated with HIV seropositivity (Appendix 8).

### PreXDR and XDR cases and associated genotypes

Next, MDR cases were further classified in preXDR and XDR cases (Figure 5). For each group of patients, the initial and final DR status was registered.

**Figure 5.**
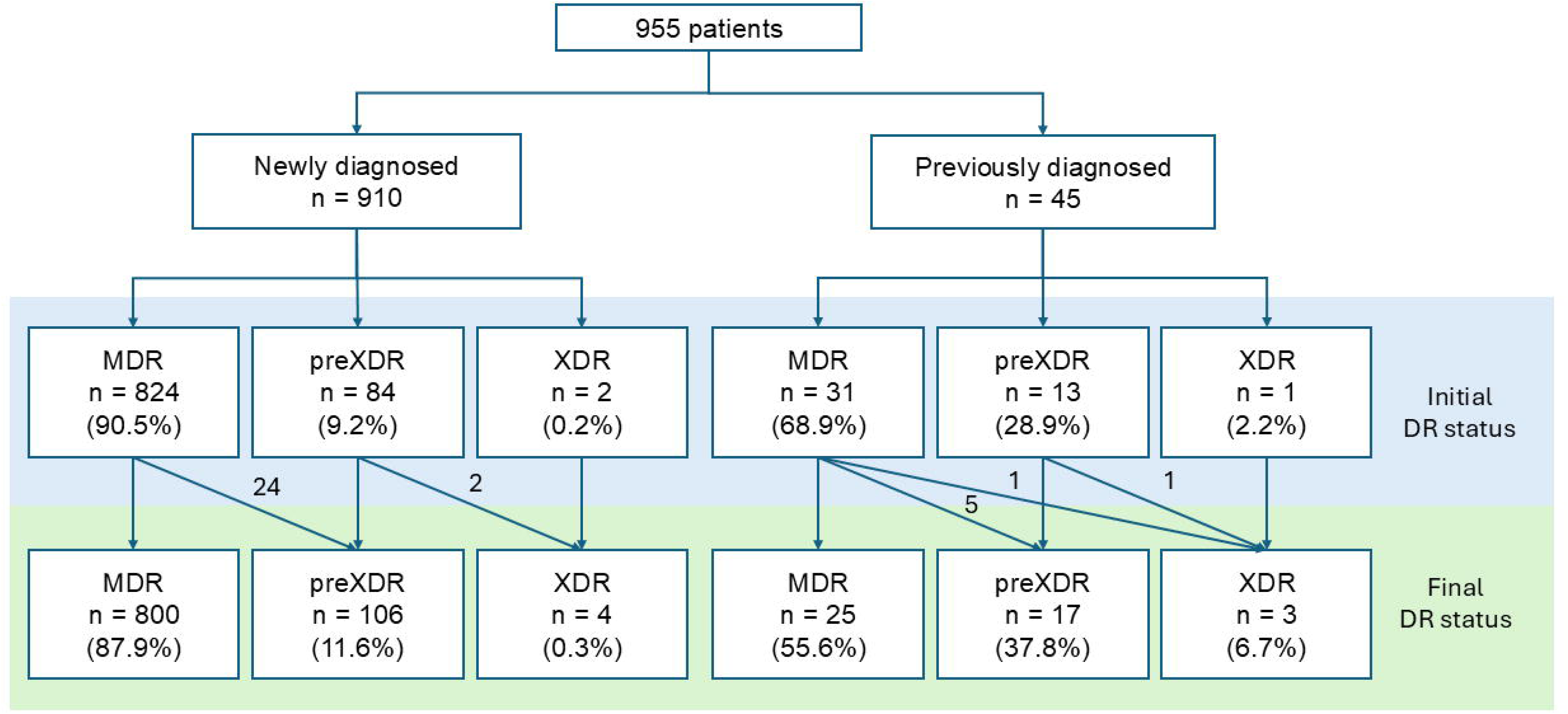
Amplification of drug resistance status of the patients, 2013 – 2022, Argentina. The cases were classified in MDR, preXDR and XDR cases according to their drug-resistance profiles to second-line drugs, among patients newly diagnosed as MDR during the study period and those who had been diagnosed in previous years. The number of cases and their percentages are indicated inside the boxes. In both groups of patients, we considered the first (initial drug-resistance status) and last (final drug-resistance status) isolates that were received during the study period. The same initial and final DR status was considered for those patients who had a single isolate. For those patients that had been diagnosed with MDR-tuberculosis in previous years, only the isolates received during the study period were considered. The number of cases that acquired a different final drug-resistance status are indicated over the arrows.

Among the new MDR-tuberculosis cases of the study period, 84 (9.2%) were preXDR upon diagnosis, and 24 acquired resistance to FLQ during their treatments (Figure 5). Most of the primary preXDR cases were residents of Buenos Aires area (65.5%) or the Northwestern provinces (25.1%). Of note, most preXDR isolates were susceptible to high-dose moxifloxacin (85/117 isolates tested, 72.6%).

The major strains only account for 21.5% of them, among which Callao2 was the most represented (11.9%). The remaining isolates were distributed across orphan genotypes and 14 minor clusters, and among these, the genotypes known as Fv and Ñ, both circulating in Buenos Aires area, were the most represented (8.3 and 7.1% respectively). In addition, we identified two primary XDR cases. The first case was due to an orphan genotype with resistance to BDQ. The patient was an young aduld female resident of a semirural area diagnosed in 2022, and had no previous treatment record, known contact with XDR cases, risk factors or travel records. The second case was due to the Rb strain, from a male healthcare worker who was diagnosed in 2013. Two cases from 2008 and 2011 were closely related (0 and 1 SNP distance respectively) and shared the *gyr*A_D94G substitution, and the 2011 and 2013 cases shared the *rpl*C_C154R mutation (associated with resistance to LNZ) (25).

Five additional cases were classified as XDR according to their final DR status (Figure 5). Two belonged to an MDR outbreak that we described recently (12) and the remaining three cases had been diagnosed in 2003, 2008 and 2011 respectively and had multiple incomplete treatments and relapses. The amplification of resistance in the last patient was described in (32), who likely became the index case of several preXDR cases due to the Ñ strain described above (data not shown). The presence of resistance mutations for the group A drugs was coincident with their phenotypic resistance.

## Discussion

Although technological advancements improved our knowledge about the prevalence of MDR genotypes, harmonizing the different outputs over extended surveillance periods poses a major challenge. Herein we integrated the results of nearly three decades of molecular surveillance of MDR-tuberculosis in Argentina and valuable insights were earned. We observed a high proportion of clustered cases, even though their relatedness is probably overestimated as discussed below. Nevertheless, clustered cases were associated with risk groups that, in turn, were overrepresented among the patients infected with the four most important clusters. Cases due to orphan genotypes represented less than 25%, and were associated with higher age, poor adherence to treatment and foreign-born patients, suggesting that they could correspond to acquired resistance cases or imported cases. Most clustered cases belonged to four strains, and three of them have been circulating in Argentina for decades (4). However, the M strain, which caused the largest documented MDR outbreak (33), started to decline. Noticeably, these patients were associated with previous treatment failure or relapse. The Rb strain was originally identified in Rosario city (4) but after the 2001–2004 outbreak among transgender women (5), most cases were found in the metropolitan area of Buenos Aires. Although the relative number of cases due to this strain remained stable during the study period, a substantial increase was observed compared to a previous study (from 4.8% in 2003 – 2009 to 14.3%) (6). In addition, this genotype became geographically widespread. Cases due to the Rb strain were strongly associated with persons living with HIV, transgender women, and with foreign-born persons. Their relationship with internal migration remains to be determined. The number of cases due to the Ra strain remained relatively stable and was chosen as reference due to its apparent epidemiological neutrality, but this could be due to under record, and specific risk groups need to be determined. Preliminary WGS-based analysis suggested that Callao2 strain could have been actively transmitted in Argentina. This strain has already been associated with transnational transmission among prison inmates that were transferred from Peru to Spain (19). Five possible active transmission chains were detected. Unfortunately, the exact transmission site of the largest cluster (Figure 4, cluster E), genomically intimately related, remains unknown. A possible relationship with the textile industry, usually associated with crowded working and living conditions, along with the temporality of the upsurge of this genotype warrants deeper epidemiological investigation. Phylogeographic analysis could shed light on the relative roles of local transmission and importation of the Callao2 cases identified in Argentina. Patients infected with the Callao2 strain were also associated with migrants born in countries other than Peru, mainly from Bolivia (11/17, 65%; data not shown), a country with relatively low proportion of MDR cases, suggesting that these patients could have been exposed in Argentina. The extended local transmission of a recently imported clone would represent a novel phenomenon in our epidemiological landscape (34).

Despite their nosocomial origin (4,7), the M, Ra and Rb strains spread into the community as their outbreaks progressed (6,35). In the current study period, cases associated with institutions such as persons deprived of liberty or health care workers were uncommon, suggesting that transmission could be occurring mainly in the community. A deeper investigation of the societal connections underlying community transmission, including the Callao2 outbreak, should be prompted.

Before 2021, XDR tuberculosis was defined according to the resistance to second-line injectable drugs and a to a FLQ (26), and used to represent 6.7% of the MDR cases in Argentina (6). The M strain is constitutively resistant to second-line injectable drugs and used to represent 40% of the XDR cases by the old definition (6). With the current definition, XDR cases were 0.7% of the all the MDR cases included in the study. The absence of dominant clones with mutations that could be missed by molecular diagnostic platforms (36) or with constitutive resistance to FLQ and group A drugs, along with the availability of all-oral second-line regimens including next-generation drugs would constitute an encouraging landscape for the control of MDR-tuberculosis compared to the previous decades. However, close monitoring of these cases is mandatory, as resistance can evolve quickly (12,35,40).

This study has important limitations. The cases included in this study represented an average of 80% of all the notified cases (Appendix 1). The real magnitude of missing cases is difficult to estimate but under-notification and under-diagnosis certainly exist, especially considering that drug-susceptibility testing is not universally performed in our country. Our sample could be biased towards most complex cases, urban centers or well-recognized risk groups, and this could be, in part, related to referral diagnosis practices. In addition, some epidemiologically relevant information could be underdeclared or biased. The higher completeness of HIV status among transgender women (82%), the apparent epidemiological neutrality of the Ra strain and the identification of a primary XDR case described herein, clearly illustrates these biases and under-sampling. The coordination among notifying agents, the referential laboratory and the administration of second-line treatments to MDR patients substantially improved in the latest years but further progress in case finding and thoroughness of epidemiological records should be prompted. Regarding our genotyping strategy, we prioritized a harmonized classification among the different methods leading to an overestimation of clustering compared to recent purely WGS-based analysis using consensus thresholds. This was the case for the O strain, which can be divided in two clades based on their spoligotype (6) but probably includes at least four different clades based on WGS analysis (Appendix 1 and 2). Transmission has been pointed as the leading driving force for the persistence of MDR-tuberculosis (20,37,38). Although our approach precludes a direct association of the observed numbers of MDR cases with recent transmission, sets the grounds for future studies that will shed light into the relative importance of transmission, reactivation, resistance acquisition and importation in our population.

## Conclusion

The results presented herein constitute important groundwork for a full transition to WGS-based surveillance, which should closely monitor the groups at higher risk identified herein. We should also bear in mind that dynamic changes can occur in relatively short timescales. The management of MDR tuberculosis in Argentina would benefit from current rapid diagnostic methods and all-oral regimes, along with a close and active monitoring of possible transmission chains and treatment follow-up. Multidisciplinary and multi-sectorial interactions would allow the full leveraging of WGS-based surveillance to translate into meaningful public health actions.

## Ethics statement

The study was conducted following the Helsinki Declaration of the World Medial Association (2013) and local regulations. The Committee in Research Ethics of the INE “Dr. Jara”, ANLIS, Argentina approved the project and waived the informed consent requirement (project code YOKOBORI05/2022).

## Data availability

Raw sequences were deposited in the NCBI or ENA. Accession numbers are available in Appendix 7. Accompanying data will be made available upon reasonable request to the corresponding author.

## Authors’ contributions

NKY conceived the study. NKY and BL designed the study. FL, RP and NKY performed the bioinformatic analysis of the genomes while LPL and DGV contributed with a refined analysis of some genomes. LPL and DGV designed the TRAP. RP, JM and IW performed the TRAP and MIRU-VNTR analysis. Genotyping and WGS data were curated and analyzed by FL, RP, JM and NKY with the contribution of LPL and DGV. NM performed second-line drug-susceptibility testing. FL and EM compiled and EM and NKY analyzed the epidemiological data. MM, AG and DP contributed with additional clinical and epidemiological data. HF compiled and analyzed WHO estimations vs. notified and referred cases. LEI and MAR performed the statistical analysis. DGV and NS contributed materials and other resources. NKY integrated the results, conceptualized them and wrote the manuscript. BL and DGV refined the first draft and suggested additional analysis. NS, RP, LPL, JM, BL, DGV and DP contributed with the final version. All the authors revised and approved the final version of the manuscript.

## Acknowledgements

We thank Javier Agüera for technical assistance in the analysis of MIRU-VNTR patterns and the “Unidad Operativa Centro Nacional de Genómica y Bioinformática” (UOCNGyB) for performing the wet lab of WGS. We also acknowledge the National Laboratory Network for the Diagnosis of Tuberculosis for their commitment and contribution to surveillance.

