## Appendix1 for "Molecular surveillance of multidrug-resistant tuberculosis at the dawn of the genomic era, Argentina, 2013–2022"

### Appendix 1. Supplementary methods

#### Representativeness of cases

In Argentina, genotyping of MDR *Mycobacterium tuberculosis* has been implemented in mid-1990’s. MDR-tuberculosis has been systematically surveyed since 2003, and as a best practice, it is highly encouraged that all the isolates detected countrywide by the National Laboratory Network for the diagnosis of tuberculosis are sent to the *Servicio de Micobacterias*, *Departamento de Bacteriología*, INEI, ANLIS for second-line drug-susceptibility testing and genotyping at referral level. MDR isolates are stored at -80°C and are available for re-culture. The number of MDR cases included is comparable to the number of cases reported in the national surveillance system (an average of 80% of the notified cases), which were, in turn, within the estimations of WHO (Supplementary table 1.1). However, these estimations have high uncertainty.

| **Supplementary table 1.1.** Estimated and notified MDR/RR tuberculosis cases in Argentina | | | | | | | | | | |
| --- | --- | --- | --- | --- | --- | --- | --- | --- | --- | --- |
|  | **WHO estimations** | | |  | **National surveillance system** | | |  | **National reference laboratory** | |
| **Year** | **Number of estimated MDR/RR cases** | **Uncertainty intervals** | |  | **Number of notified MDR/RR cases*** | **Coverage vs. WHO** | **Number of MDR cases*** |  | **Number of MDR cases*** | **Coverage vs. notification** |
|  |  | **low** | **high** |  |  |  |  |  |  |  |
| **2013** | **-** | **-** | **-** |  | **-** | **-** | 80 |  | 105 | 131.3% |
| **2014** | **-** | **-** | **-** |  | **-** | **-** | 117 |  | 103 | 88.0% |
| **2015** | 310 | 78 | 540 |  | 120 | 38.7% | 93 |  | 89 | 95.7% |
| **2016** | 330 | 64 | 590 |  | 114 | 34.5% | 87 |  | 88 | 101.1% |
| **2017** | 320 | 42 | 600 |  | 121 | 37.8% | 111 |  | 90 | 81.1% |
| **2018** | 320 | 21 | 610 |  | 174 | 54.4% | 147 |  | 105 | 71.4% |
| **2019** | 347 | 0 | 699 |  | 229 | 66.0% | 169 |  | 106 | 62.7% |
| **2020** | 277 | 0 | 578 |  | 132 | 47.6% | 91 |  | 82 | 90.1% |
| **2021** | 348 | 0 | 753 |  | 175 | 50.3% | 124 |  | 114 | 91.9% |
| **2022** | 356 | 0 | 803 |  | 222 | 62.3% | 131 |  | 89 | 67.9% |
| **Mean** | 326 | 26 | 647 |  | 161 | 48.9% | 119 |  | 95 | 80.1% |
| Source: WHO and National surveillance program for tuberculosis, Argentina. WHO estimates were retrived from the Global Tuberculosis Report 2025, application version (July 2025) which did not include data from 2013 and 2014. *Includes incient and previously diagnosed cases. | | | | | | | | | | |

#### Combined genotyping strategy

Our laboratory keeps a genotyping database of MDR isolates evaluated with *IS*6110-RFLP and spoligotyping (1998-2010, ~1400 isolates), 15 loci MIRU-VNTR (2012-2020, ~1050 isolates) or WGS (2021 onwards, ~400 isolates). Some older isolates have been analyzed by WGS in prior research projects. Targeted regional allele-specific oligonucleotide PCRs (TRAP) is a multiplexed in-house PCR designed to detect specific SNPs of *M. tuberculosis* clones of concern to confirm or discard their identity (1–3). Currently, three TRAPs directed to the detection of M, Ra and Rb strains, the most prevalent MDR clones in Argentina in the previous period, are available. Part of the isolates received during 2015 – 2020 were identified using TRAP, and MIRU-VNTR was applied to non-M, non-Ra and non-Rb isolates. Partial genotyping analysis based on TRAP-M and TRAP-Ra were published elsewhere (2). Validation assays were performed in each methodological transition to ensure the continuity of data, with a special focus in the clusters identified by each technique. Correspondence between MIRU-VNTR15 and TRAP has been previously published (2,3).

For this study, a consolidated classification method was applied for the assignment of the genotype of each isolate. Correspondence between MIRU-VNTR15 and TRAP has been previously published (2,3). To define the WGS-based clusters, a cutoff value of ≤20 SNP was chosen. Their phylogenetic relatedness was checked. This threshold is higher than international standards that are usually adopted in purely WGS-based analysis (4–6). However, our main goal was to ensure concordance with MIRU-VNTR-based classification, which was verified for the most important clusters. Representative isolates of the most important MDR clusters had been sequenced in the past (3,7–9). Clusters that were newly identified along with orphan isolates detected by WGS were evaluated with MIRU-VNTR and were classified following the above-mentioned criteria. We performed a sensitivity and specificity analysis comparing MIRU-VNTR15 and WGS for the isolates from 2018 which had been typed using both methods, as detailed in Appendix 2. For 15 loci MIRU-VNTR-based classification, a single difference in one VNTR was exceptionally tolerated when strong epidemiological and/or microbiological data supported their relatedness. For the remaining isolates, a cluster was defined when at least two isolates from different patients shared an identical MIRU-VNTR pattern or had a SNP distance ≤20, supported by phylogenetic relationships. The O strain pattern corresponded to at least four WGS-defined clusters based on a 20 SNP threshold. Two of these variants had been characterized in the past (10). Because many O strain isolates were exclusively evaluated by MIRU-VNTR (50.61%), we chose to consider these isolates as a single genotype for this study. Further research is warranted to have a clearer insight into the O genotype and its diversity. Based on these observations, we evaluated the correspondence between MIRU-VNTR considering: 1) raw data; 2) collapsing isolates with a single difference in MIRU-VNTR15 type, with epidemiological or microbiological evidence supporting their relatedness, in the same cluster as the parental strain (column MIRU-VNTR-collapsed vs. WGS in Supplementary table 1.2) and 3) collapsing all the O strain subclades identified by WGS. The latter strategy led to an agreement of 96.08% (Supplementary table 1.2). Two cases were probably misclassified by MIRU-VNTR, and the WGS-based genotype was considered for the final analysis included in the manuscript. This strategy has several limitations, including probable overestimation of clustering due to loose criteria and possible misclassification of small clusters for which complementary information is scarce.

| **Supplementary table 1.2. Sensitivity and specificity analysis comparing MIRU-VNTR and WGS.** | | | |
| --- | --- | --- | --- |
|  | **MIRU-VNTR**  **vs. WGS** | **MIRU-VNTR collapsed**  **vs. WGS** | **MIRU-VNTR collapsed  vs. WGS collapsed** |
| **Macro average (considering clusters)** | |  |  |
| Sensitivity | 0.8000 | 0.9333 | 0.9809 |
| Specificity | 0.9951 | 0.9988 | 0.9988 |
| **Micro average (considering clusters and n per cluster)** | | |  |
| Sensitivity | 0.7647 | 0.8431 | 0.9608 |
| Specificity | 0.9951 | 0.9988 | 0.9988 |
| **Kohen's kappa** | 0.748 | 0.831 | 0.957 |
| p-value | <0.001 | <0.001 | <0.001 |
| Agreement (%) | 76.47% | 84.31% | 96.08% |
| MIRU-VNTR collapsed: isolates with a single difference in MIRU-VNTR15 type, with epidemiological or microbiological evidence supporting their relatedness, were considered in the same cluster as the parental strain (number of isolates for M, Tu and O strains: 2, 2 and 1 respectively).  WGS collapsed: O strain variants (Ob, Os, Or and O_other) were collapsed into a single category (O).  Raw data and detailed analysis can be found in Appendix 2. | | | |
