## Appendix4 for "Molecular surveillance of multidrug-resistant tuberculosis at the dawn of the genomic era, Argentina, 2013–2022"

### Appendix 4. Multivariate analysis

To determine which combination of demographic and clinical variables significantly differentiated the patients infected with the different groups of patients (clustered vs. orphan cases and the cases due to the four major strains), a multivariate analysis (multinomial logistic regression) was carried out with a backward stepwise selection mechanism. We started with a saturated model and in each step the model is analyzed, eliminating those variables that led to a lower AIC (Akaike Information Criterion) upon removal. The process was stopped when no removal improved the AIC. A P value <0.05 and 95% confidence intervals (CI) that did not cross 1 were considered significant.

The categorical variables with more than two categories were recodified as binary indicators (yes/no) for their inclusion in the model, which allows the estimation of independent effects of each category compared to a reference, controlled by the other covariables of the model. Gender was separated in two indicators: cis women (yes/no) and trans women (yes/no), taking cis men as the reference category. Country of birth was also separated in two indicators: born in Peru (yes/no) and other foreign born (yes/no), taking born in Argentina as the reference category.

#### Clustered vs. orphan cases (Table 1)

**Dependent variable.** Clustering status with two categories (clustered and orphan).

**Independent variables.** Age (per year), Gender (three categories: cisgender men, cisgender women and transgender women), country of birth (three categories: Argentina, Peru, other foreign), HIV serology (three categories: positive, negative and unknown), poor adherence (binary), known contact (binary), treatment failure (binary), people deprived of liberty (binary), substance abuse (binary), diabetes (binary), alcoholism (binary), health care worker (binary), and smoking (binary).

**Number of cases.** 863 cases (208 orphan, 655 clustered)

**Final model:**

Clustering **=** Age + cisgender women (cisgender male = reference) + transgender woman (cisgender male = reference) + country of birth Peru (Argentina = reference) + country of birth other foreign (Argentina = reference) + HIV positive (negative = reference) + poor adherence + known contact + treatment failure + people deprived of liberty + substance abuse + diabetes

#### Major strains (M, Ra, Rb and Callao2; Table 3)

**Dependent variable.** Strain with four categories of interest (M, Ra, Rb, Callao2). The Ra strain was selected as the reference value, due to its higher n. It is also the strain without extreme specific associations, which makes it the most epidemiologically neutral point of comparison.

**Independent variables**

| **Supplementary table 4.1.** Epidemiological variables or the patients infected with the four major strains, descriptive statistics and variable inclusion/inclusion for multivariate analysis | | | | | |
| --- | --- | --- | --- | --- | --- |
| **Variable** | **Ra (reference)**  **n = 119** | **M**  **n = 75** | **Rb**  **n = 128** | **Callao2**  **n = 90** | **Variable selection** |
| Age (years [IQR]) | 36 [26-45.2] | 37 [30-48] | 35 [28-43] | 29 [21-41]‡ | Included |
| Gender |  |  |  |  |  |
| Cis men (reference) | 71 (59.7%) | 44 (58.7%) | 80 (62.5%) | 56 (62.2%) |  |
| Cis women | 47 (39.5%) | 31 (41.3%) | 19 (14.8%) | 31 (34.4%) | Excluded (stepwise) |
| Trans women | 1 (0.8%) | 0 (0%) | 29 (22.7%) | 3 (3.3%) | Included |
| Country of birth* |  |  |  |  |  |
| Argentina (reference) | 103 (85.8%) | 71 (93.4%) | 100 (77.5%) | 35 (38.9%) |  |
| Peru | 1 (0.8%) | 0 (%) | 8 (6.2%) | 38 (42.2%) | Included |
| Other foreign | 16 (13.4%) | 5 (6.6%) | 21 (16.3%) | 17 (18.9%) | Included |
| HIV |  |  |  |  |  |
| Positive | 18 (15%) | 15 (19.7%) | 63 (48.8%) | 11 (12.2%) | Included |
| Other | 102 (85%) | 61 (80.3%) | 66 (51.2%) | 79 (87.8%) |  |
| Poor adherence† | 5 (4.2%) | 10 (13.2%) | 15 (11.6%) | 4 (4.4%) | Included |
| Treatment failure† | 6 (5%) | 13 (17.1%) | 9 (7.0%) | 13 (14.4%) | Included |
| Diabetes† | 10 (8.3%) | 5 (6.6%) | 4 (3.1%) | 1 (1.1%) | Excluded (stepwise) |
| Substance abuse† | 3 (2.5%) | 2 (2.6%) | 4 (3.1%) | 6 (6.7%) | Excluded (n<10) |
| Alcoholism† | 1 (0.8%) | 2 (2.6%) | 3 (2.3%) | 4 (4.4%) | Excluded (n<10) |
| Health care workers† | 1 (0.8%) | 4 (5.3%) | 3 (2.3%) | 0 (0%) | Excluded (n<10) |
| Smoking† | 1 (0.8%) | 0 (0%) | 2 (1.6%) | 1 (1.1%) | Excluded (n<10) |
| Known contact† | 8 (6.7%) | 15 (19.7%) | 10 (7.7%) | 21 (23.3%) | Excluded (n<10) |
| PDL† | 6 (5%) | 11 (14.5%) | 3 (2.3%) | 3 (3.3%) | Excluded (n<10) |
| *Among the 14 patients classified as “other foreign”, the country of birth was unknown in 3 cases.  †Binary variables were classified in known/unknown. | | | | | |

**Number of cases.** Of the 412 cases due to the four most important strains, 391 were included due to completeness of the selected variables.

**Initial model:**

Strain **=** Age + cisgender women (cisgender male = reference) + transgender woman (cisgender male = reference) + country of birth Peru (Argentina = reference) + country of birth other foreign (Argentina = reference) + HIV positive + poor adherence + treatment failure + diabetes

**Final model:**

Strain = age + transgender women + country of birth Peru + country of birth other foreign + HIV positive + poor adherence + treatment failure

- Ra = reference category.

The stepwise model retained 7 of the 9 candidate variables. The two variables that were eliminated — female gender and diabetes — did not contribute to the discrimination among strains once adjusted for the others.
