## Appendix5 for "Molecular surveillance of multidrug-resistant tuberculosis at the dawn of the genomic era, Argentina, 2013–2022"

### Appendix 5. Lineages based on MIRU-VNTR, WGS and in silico spoligotyping, MDR *M. tuberculosis*, Argentina 2013–2022

Lineages based on MIRU-VNTR and WGS were determined, based on their availability. For the isolates analyzed by WGS, in silico spoligotypes were obtained. Isolates from lineages different from Euro-American/lineage 4 were extremely rare in our context.

| **Supplementary table 5.1.** MIRU-VNTR-based lineage assignment | | | | |
| --- | --- | --- | --- | --- |
| **Lineage** | **Clusters** | | **Orphan** | |
|  | **n*** | **%** | **n*** | **%** |
| Euro-American | 44 | 97.78 | 175 | 97.76 |
| East-Asian | 1† | 2.22 | 3‡ | 1.68 |
| East-African-Indian | 0 | 0 | 1§ | 0.56 |
| Total | 45 |  | 179 |  |
| *Number of genotypes. †Part of a minor cluster that involved three cases including patients diagnosed before 2013. Two patients were born in Peru, and the remaining patient was a patient born in Argentina who was deprived of liberty at the time of diagnosis. ‡These patients were foreign-born (one in Ukraine and two in Peru). §The patient was resident of Chaco province which is close to the Argentina-Paraguay-Brazil triple border. This lineage is extremely rare in our country and this result should be checked with more precise methods.  Run TB-lineage was used to infer the MIRU-VNTR-based genetic lineage (Shabbeer et al., 2012). | | | | |

| **Supplementary table 5.2.** WGS-based lineage assignment | | |
| --- | --- | --- |
| **Lineage** | **Clusters** | **Orphan** |
| lineage4 | 34 (100%) | 62 (93.9%) |
| lineage4.1.1 | 3 | 2 |
| lineage4.1.1.3 | 1 | 3 |
| lineage4.1.2 | 1 | 3 |
| **lineage4.1.2.1** | **6** | **11** |
| lineage4.1.2.1.1 | 1 | 1 |
| lineage4.1.4 | 0 | 1 |
| **lineage4.3.2** | **5** | **7** |
| **lineage4.3.3** | **7** | **12** |
| lineage4.3.4.1 | 1 | 0 |
| lineage4.3.4.2 | 2 | 3 |
| lineage4.4.1.1 | 1 | 4 |
| lineage4.7 | 2 | 3 |
| **lineage4.8** | 2 | **7** |
| lineage4.9 | 0 | 1 |
| lineage4 | 2 | 4 |
| lineage2 | 0 (0%) | 4 (6.1%) |
| **lineage2.2.1** | 0 | **3*** |
| lineage2.2.2 | 0 | 1† |
| Total | 34 | 66 |
| *One of the patients was born in Peru. The nationality of the remaining patients were unknown. †The patient was born in Corea.  Number of genotypes and percentages among the isolates analyzed by WGS. TB-profiler was used to infer the lineage (Phelan, 2023). The most prevalent sublineages are highlighted in bold face. | | |

| **Supplementary table 5.3.** WGS-based in silico spoligotypes | | | |
| --- | --- | --- | --- |
| **Spoligoyping clade** | **SIT** | **Clusters** | **Orphan** |
| BEIJING | 1 | 0 (0%) | **4 (6.06**%**)** |
| H1 | 47 | 1 | 0 |
|  | 62 | 0 | 1 |
|  | 531 | 1 | 0 |
|  | Subtotal | 2 (5.41%) | 1 (1.52%) |
| H2 | 2 | 1 (2.70%) | 1 (1.52%) |
| LAM1 | 20 | 1 | 0 |
|  | 469 | 0 | 1 |
|  | Subtotal | 1 (2.70%) | 1 (1.52%) |
| LAM3 | 33 | **3** | **3** |
|  | 130 | 1 | 1 |
|  | 211 | 1 | 0 |
|  | 1537 | 0 | 1 |
|  | 2626 | 0 | 1 |
|  | Unknown | 1 | 0 |
|  | Subtotal | 6 (16.22%) | 6 (9.09%) |
| LAM4 | 60 | 0 (0%) | 1 (1.52%) |
| LAM5 | 93 | 2 | 1 |
|  | 136 | 0 | 1 |
|  | 725 | 1 | 1 |
|  | Subtotal | 3 (8.11%) | 3 (4.55%) |
| LAM6 | 1355 | 1 (2.70%) | 0 (0%) |
| LAM9 | 42 | 2 | **7** |
|  | 2263 | 1 | 0 |
|  | 2331 | 1 | 1 |
|  | Subtotal | 4 (10.81%) | 8 (12.12%) |
| S | 34 | 0 | 2 |
|  | 71 | 0 | 1 |
|  | Subtotal | 0 (0%) | 3 (4.55%) |
| T1 | 53 | **5** | **11** |
|  | 159 | **3** | 1 |
|  | 219 | 0 | 1 |
|  | 222 | 0 | 1 |
|  | 253 | 0 | 1 |
|  | 281 | 0 | 1 |
|  | 291 | 0 | 2 |
|  | 373 | 1 | 1 |
|  | 1105 | 0 | 1 |
|  | 1122 | 0 | 1 |
| (continued) | Unknown | 0 | 1 |
|  | Subtotal | 9 (24.32%) | 22 (33.33%) |
| T3 | 37 | 1 | 2 |
|  | Unknown | 0 | 1 |
|  | Subtotal | 1 (2.70%) | 3 (4.55%) |
| T4 | 40 | 0 (0%) | 1 (1.52%) |
| T5-Madrid2 | 58 | 1 (2.70%) | 3 (4.55%) |
| X1 | 119 | 1 | 1 |
|  | 1080 | 0 | 1 |
|  | Subtotal | 1 (2.70%) | 2 (3.03%) |
| X3 | 91 | 1 | 1 |
|  | 92 | 1 | 0 |
|  | Subtotal | 2 (5.41%) | 1 (1.52%) |
| Unknown | 4 | 1 | 0 |
|  | 450 | 1 | 2 |
|  | 881 | 0 | 1 |
|  | F4 | 1 | 0 |
|  | Unknown | 2 | 3 |
|  | Subtotal | 5 (13.51%) | 6 (9.09%) |
| **Total** |  | **37*** | **66** |
| *Three clusters, including the O strain, had more than one spoligotype.  N of genotypes based on WGS-based *in silico* spoligotyping using TB-profiler (Phelan, 2023). Number of genotypes by clade and SIT are shown. The percentage represented by each clade among clustered and Orphan genotypes is shown between brackets. The most prevalent SITs are indicated in bold face. Five spoligotypes were orphan types. | | | |
