## Appendix6 for "Molecular surveillance of multidrug-resistant tuberculosis at the dawn of the genomic era, Argentina, 2013–2022"

### Appendix 6. Cluster sizes and cumulative number of newly diagnosed cases of the four major MDR *M. tuberculosis* clusters, Argentina, 2013–2022

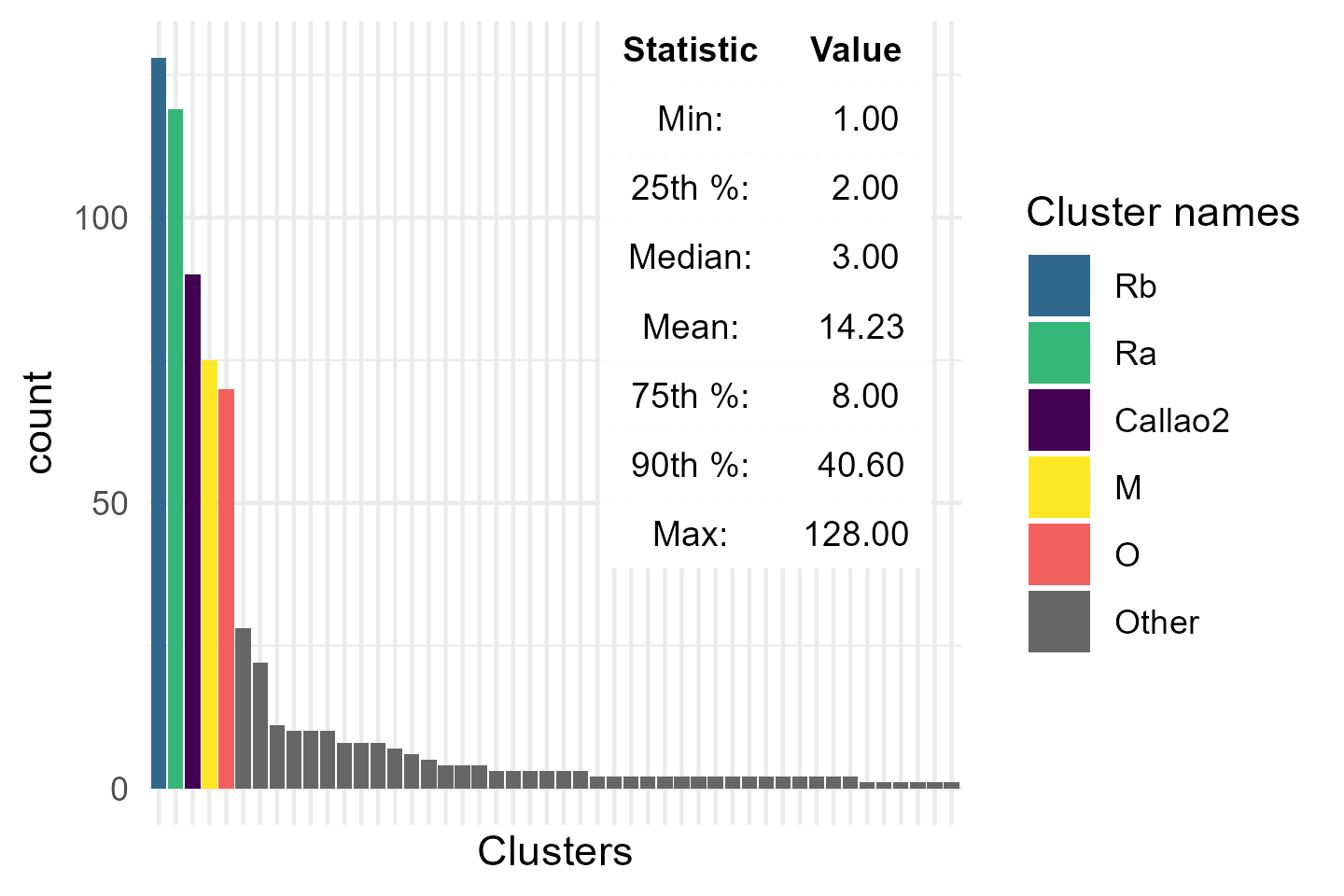

**Supplementary figure 6.1.** Histogram shows the number of isolates from newly diagnosed cases per cluster. The table in the inset shows the descriptive statistic values for the number of isolates per cluster. The clusters belonging to the top decile, namely the Rb, Ra, Callao2, M and O strains, are highlighted in colors.

A total of 48 clusters were identified. Among them, five clusters belonged to the top decile according to their cluster sizes. After excluding the O strain, which most likely represents several diverse clusters (see the main text and Appendix 1), the remaining four clusters were selected as the major clusters of the period for further analysis.
