## Appendix8 for "Molecular surveillance of multidrug-resistant tuberculosis at the dawn of the genomic era, Argentina, 2013–2022"

### Appendix 8. Risk factors associated with minor and major MDR-*M. tuberculosis* clusters, Argentina, 2013–2022

| **Supplementary Table 8.1** | | | | |
| --- | --- | --- | --- | --- |
| **Risk factor** | **Minor clusters** | **Major clusters** | **Odds ratio**  **[95% CI]** | **p value** |
| Declared contact with an MDR patient (%)* | 52/201 (25.9 %) | 54/412 (13.1 %) | 2.3 [1.5–3.6] | <0.001 |
| HIV seropositivity (%) | 21/201 (10.4 %) | 104/414 (25.2 %) | 0.3 [0.2–0.6] | <0.001 |
| *Only four of the declared contacts were not family members.  CI: confidence interval.  The O strain was excluded from this analysis because it probably includes several strains that are indistinguishable by MIRU-VNTR (Appendix 1). Other risk factors were not associated with cluster size. Chi-squared test was used. Odds ratio and CIs were calculated using the Fishers’ method. | | | | |
